# Divergent Eosinophilic and Paradoxical Inflammatory Adverse-Event Signatures Across Five Type 2 Asthma Biologics: A FAERS Active-Comparator Disproportionality Analysis

**DOI:** 10.64898/2026.09.21.26363575

**Authors:** Adalia Cristiana Iancu, Marwan Kobtan, Nada Soliman, Shahinazhanem Abdellatif

**Affiliations:** Carol Davila University of Medicine and Pharmacy, Bucharest, Romania; Victor Babes University of Medicine and Pharmacy, Bucharest, Romania; Cairo University Faculty of Medicine, Kasr Al Ainy, Cairo, Egypt; New Giza University, Giza, Egypt; University Canada West, Vancouver, BC, Canada

**Keywords:** pharmacovigilance, disproportionality analysis, FAERS, reporting odds ratio, type 2 inflammation, biologics, asthma, eosinophilic granulomatosis with polyangiitis, dupilumab, mepolizumab, benralizumab, tezepelumab, omalizumab

## Abstract

**Background:** Five biologics target sequential nodes of the type 2 (T2) inflammatory axis in severe asthma and have each been anecdotally linked to eosinophilic or paradoxical inflammatory adverse events. Whether these reflect a class effect or drug-specific signatures is untested.

**Methods:** We performed a case-noncase disproportionality analysis of FAERS, 2022–2024. Asthma-indication reports naming tezepelumab, dupilumab, mepolizumab, benralizumab, or omalizumab as primary suspect were compared against the pooled other four (active-comparator design) across 7 pre-specified eosinophilic, vasculitic (including EGPA), cutaneous, and ocular outcomes. A signal was robust if the reporting odds ratio (ROR) 95% CI lower bound exceeded 1 and a Bayesian criterion was also met.

**Results:** Among 50,360 reports (dupilumab n=29,652; mepolizumab n=8,846; omalizumab n=5,484; benralizumab n=4,372; tezepelumab n=2,006), benralizumab showed robust signals for EGPA (ROR 5.06), vasculitis (ROR 4.60), and peripheral eosinophilia (ROR 1.80). Mepolizumab showed a robust vasculitis signal (ROR 1.85); its similarly sized EGPA association narrowly missed the robustness criterion despite significance after correction for multiple testing. Dupilumab showed no EGPA/vasculitis excess but a robust ocular-inflammation signal (ROR 7.81). Tezepelumab and omalizumab showed no robust signal in any domain. EGPA/vasculitis onset occurred earlier after dupilumab (median 20–44 days) than anti-IL-5-axis agents (median 195–550 days; p≤0.012); mepolizumab’s signal did not reproduce over the full 2015–2024 window (calendar-time confounding).

**Conclusion:** FAERS active-comparator analysis reveals drug-specific, not class-uniform, adverse-event reporting patterns: anti-IL-5-axis agents disproportionately report EGPA/vasculitis, and dupilumab disproportionately reports ocular inflammation with markedly earlier onset. These hypothesis-generating signals cannot establish causality and require confirmation through case-level or comparative pharmacoepidemiological studies.

## 1. Introduction

Type 2 (T2) inflammation drives the eosinophilic, immunoglobulin E (IgE)-associated endotype underlying most severe asthma. Five biologics now target this axis at sequential nodes. Tezepelumab neutralizes the epithelial alarmin thymic stromal lymphopoietin (TSLP) upstream of T2-cell activation; dupilumab blocks the interleukin (IL)-4 receptor alpha (IL-4Rα) subunit shared by IL-4 and IL-13; mepolizumab neutralizes IL-5; benralizumab blocks the IL-5 receptor alpha (IL-5Rα) subunit and additionally depletes eosinophils via antibody-dependent cellular cytotoxicity; and omalizumab neutralizes circulating IgE downstream of B-cell class-switching (Figure 1). Because all five ultimately dampen the same inflammatory cascade, a shared adverse-event profile might be expected. Yet each has also been anecdotally linked to a distinct constellation of eosinophilic or paradoxical inflammatory events — peripheral eosinophilia, eosinophilic pneumonia, eosinophilic granulomatosis with polyangiitis (EGPA) and other vasculitic phenomena, paradoxical cutaneous inflammation, and ocular surface inflammation — raising the question of whether these represent a shared class effect of T2-axis blockade or mechanistically distinct, drug-specific signatures tied to each agent’s precise molecular target.

**Figure 1.**
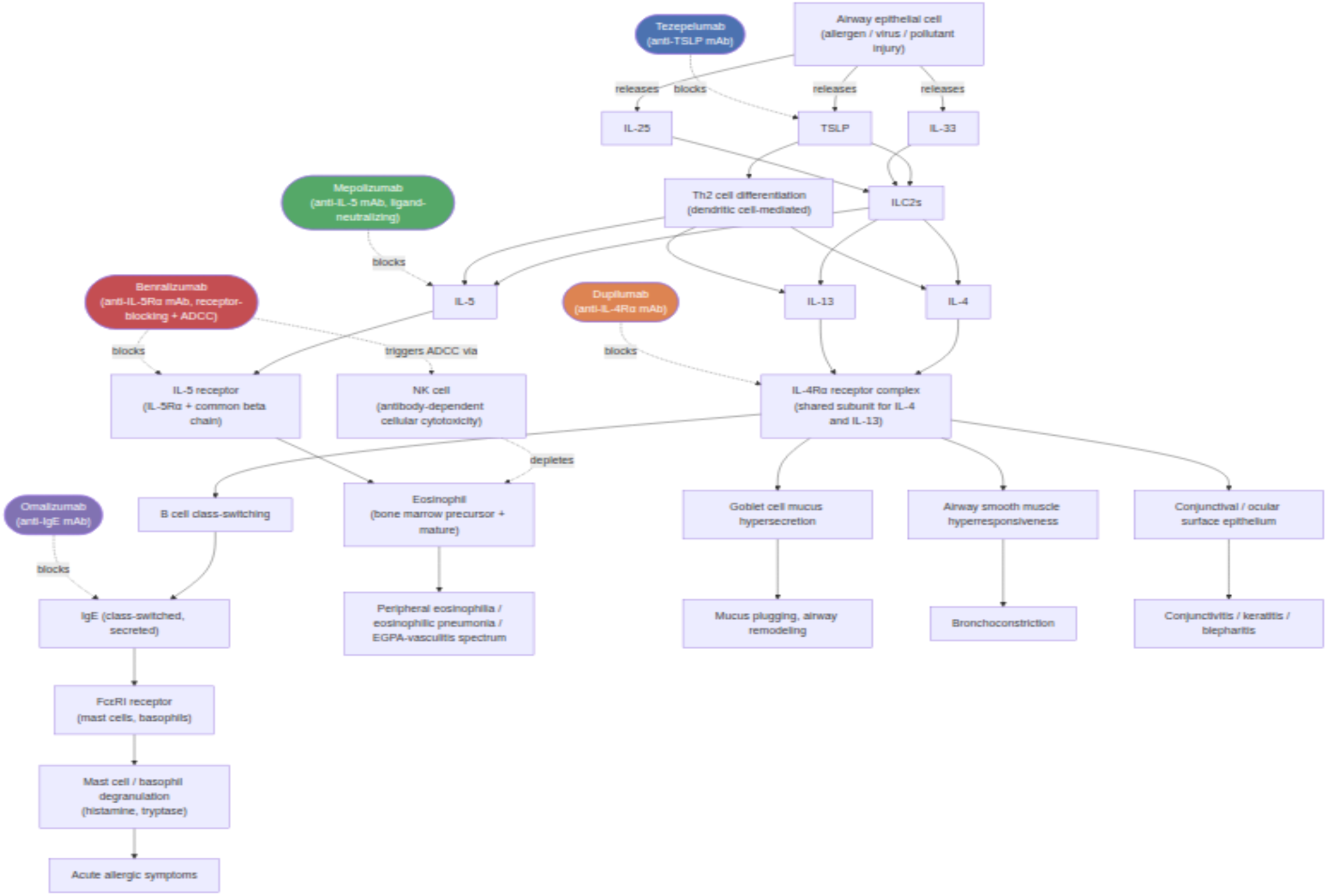
Schematic of the type 2 inflammatory cascade and each biologic’s molecular site of action. Tezepelumab neutralizes thymic stromal lymphopoietin (TSLP) upstream of type 2 cell activation; dupilumab blocks the interleukin (IL)-4 receptor alpha (IL-4Rα) subunit shared by IL-4 and IL-13; mepolizumab neutralizes IL-5; benralizumab blocks the IL-5 receptor alpha (IL-5Rα) subunit and additionally triggers antibody-dependent cellular cytotoxicity via natural killer cells; and omalizumab neutralizes circulating immunoglobulin E (IgE). This figure illustrates established pharmacological mechanism of action based on general immunopharmacology and each drug’s labeled mechanism; it does not represent measured molecular or receptor-level data from FAERS and is provided as interpretive background for the Introduction and Discussion, not as an empirical study result.

Spontaneous-report pharmacovigilance databases such as the FDA Adverse Event Reporting System (FAERS) enable post-marketing signal detection at a scale unattainable in randomized trials, which are underpowered for rare outcomes such as EGPA. However, conventional disproportionality analyses that compare a drug against the entire FAERS background are vulnerable to confounding by indication: because all five biologics are prescribed predominantly for eosinophilic, T2-high asthma, each will appear disproportionately associated with eosinophilic outcomes relative to “all of FAERS,” independent of any true between-drug difference. An active-comparator design — benchmarking each biologic against the pooled other agents used for the same indication — mitigates this confounding and has not, to our knowledge, previously been applied across all five currently marketed T2 asthma biologics simultaneously.

We therefore conducted a case-noncase disproportionality analysis of FAERS to directly compare the relative reporting of pre-specified eosinophilic and paradoxical inflammatory adverse-event domains across tezepelumab, dupilumab, mepolizumab, benralizumab, and omalizumab among asthma-indication reports from 2022 to 2024, the period during which all five agents were simultaneously marketed.

## 2. Materials and Methods

### 2.1 Study design

This was a disproportionality analysis of individual case safety reports (ICSRs) from a spontaneous adverse-event reporting database, conducted and reported in accordance with the READUS-PV guideline for disproportionality analyses using ICSRs^1^. We used a case/non-case design with an active-comparator reference group as the primary analysis (Analysis A) and a conventional whole-database background comparison as a secondary analysis (Analysis B). Four pre-specified direct pairwise comparisons between individual biologics (Supplementary Table S1) and ten pre-specified sensitivity analyses (S1–S10; Table 2) were additionally performed. No case-by-case clinical review or formal causality-assessment algorithm (e.g., WHO-UMC, Naranjo) was applied to individual ICSRs; this is therefore a signal-detection, not a signal-validation, study.

**Table 1.** Study population by biologic, primary analysis (asthma-only, Primary-Suspect-only, 2022–2024).

| Biologic | Molecular target | N (asthma cohort, PS-only) | % of total |
| --- | --- | --- | --- |
| Dupilumab | Anti-IL-4Rα | 29,652 | 58.9% |
| Mepolizumab | Anti-IL-5 | 8,846 | 17.6% |
| Omalizumab | Anti-IgE | 5,484 | 10.9% |
| Benralizumab | Anti-IL-5Rα | 4,372 | 8.7% |
| Tezepelumab | Anti-TSLP | 2,006 | 4.0% |
| **Total** | — | **50,360** | **100%** |

*PS = Primary Suspect; IL = interleukin; IL-4Rα = interleukin-4 receptor alpha; IL-5Rα = interleukin-5 receptor alpha; IgE = immunoglobulin E; TSLP = thymic stromal lymphopoietin. Total deduplicated FAERS reports in the 2022–2024 window, any drug or indication (Analysis B background): 4,451,126. Full any-indication population, any of the five biologics as Primary Suspect (Analysis B/S6 numerator): 262,191.*

**Table 2.** Pre-specified sensitivity analyses S1–S10 (primary 2022–2024 cohort unless noted).

| ID | Description | Key finding |
| --- | --- | --- |
| S1 | Common calendar, 2022–2024 | Equivalent to the primary analysis (this manuscript) |
| S2 | Full post-marketing window per drug, 2015–2024 | Benralizumab (EGPA ROR 3.08, 95% CI 2.36–4.02; vasculitis ROR 2.93, 2.32–3.71; peripheral eosinophilia ROR 2.16, 1.78–2.62) and dupilumab (ocular inflammation ROR 8.25, 6.94–9.81; eosinophilic pneumonia ROR 2.26, 1.49–3.41) signals replicated. Mepolizumab's EGPA and vasculitis signals became null ($\log_2$ ROR +0.38 and +0.28). A new omalizumab rosacea signal emerged (ROR 2.52, 1.38–4.59; not seen in the primary period; small n). |
| S3 | Primary Suspect role only | Equivalent to the primary analysis |
| S4 | Primary + Secondary Suspect roles | Cohort grew from 50,360 to 58,466.<br>Benralizumab/mepolizumab EGPA-vasculitis signals and dupilumab's ocular signal persisted; dupilumab's peripheral-eosinophilia and eosinophilic-pneumonia point estimates attenuated toward or below the null. |
| S5 | Asthma-only indication restriction | Equivalent to the primary analysis |
| S6 | All indications (no asthma restriction) | Directionally consistent with the primary analysis |
| S7 | Serious reports only | Directionally consistent with the primary analysis |
| S8 | Healthcare-professional reporters only | Directionally consistent with the primary analysis |
| S9 | United States reports only | Directionally consistent with the primary analysis |
| S10 | Exclusion of cross-biologic-exposed cases | No cross-exposed cases were identified in the primary cohort (0 of 50,360); numerically identical to the primary analysis. |

*EGPA = eosinophilic granulomatosis with polyangiitis; ROR = reporting odds ratio; CI = confidence interval.*

### 2.2 Data source, access, and pre-processing

Data were obtained from FAERS, a publicly available spontaneous-report ICSR database maintained by the FDA, which receives reports for all FDA-regulated drugs and therapeutic biological products marketed in the United States, submitted by manufacturers, healthcare professionals, and consumers, without restriction to drug class or indication. Adverse events are coded using Medical Dictionary for Regulatory Activities (MedDRA) Preferred Terms; the specific MedDRA version(s) in effect could not be confirmed from the publicly available extracts, which do not embed a per-quarter version tag.

All quarterly ASCII Data Extract Files covering January 2015 through December 2024 (40 quarters) were downloaded from FDA’s public static export endpoint on 13 September 2026. The primary analysis period was restricted to January 1, 2022 – December 31, 2024, the period during which all five study biologics were simultaneously marketed (tezepelumab was first approved December 2021); a secondary analysis repeated the design over each drug’s complete post-marketing window through December 2024.

Reports were deduplicated at the case level, retaining the highest case version and collapsing duplicate identical drug-event pairs. Reports naming two or more of the five study biologics simultaneously as suspect drugs were excluded from the active-comparator analysis (cross-exposure exclusion; sensitivity analysis S10). This procedure cannot eliminate residual duplicate reporting under distinct case identifiers, a limitation inherent to de-identified public FAERS data.

### 2.3 Study population and outcome definitions

The primary population comprised ICSRs in which one of the five study biologics was recorded as Primary Suspect and linked to an asthma-related indication on that same drug record. Reports were excluded if the same drug record was also linked to a major non-asthma indication (e.g., atopic dermatitis, chronic rhinosinusitis with nasal polyps, eosinophilic esophagitis, chronic urticaria, hypereosinophilic syndrome, or EGPA), to limit confounding by indication and reverse-causation bias, because several of these conditions — notably hypereosinophilic syndrome and EGPA — are both study outcomes and approved indications for individual anti-IL-5-axis agents.

Outcome variables comprised five pre-specified clinical domains, operationalized as seven composite or single-Preferred-Term (PT) outcome measures: (1) peripheral eosinophilic phenotype (eosinophilia, eosinophil count increased, hypereosinophilic syndrome); (2) eosinophilic pneumonia; (3a) EGPA and (3b) a broader narrow-vasculitis PT list, analyzed separately; (4a) a psoriasis-family composite and (4b) rosacea, analyzed separately; and (5) an ocular inflammatory composite (conjunctivitis, keratitis, blepharitis, dry eye, ocular rosacea). Six additional exploratory autoimmune/inflammatory PTs were assessed as a single non-primary “exploratory autoimmune (any)” composite for the network-perturbation heatmap only (Figure 2); this composite combines heterogeneous conditions dominated by non-specific PTs such as arthralgia and is reported descriptively, excluded from the primary hypothesis set and from formal robust-signal classification.

**Figure 2.**
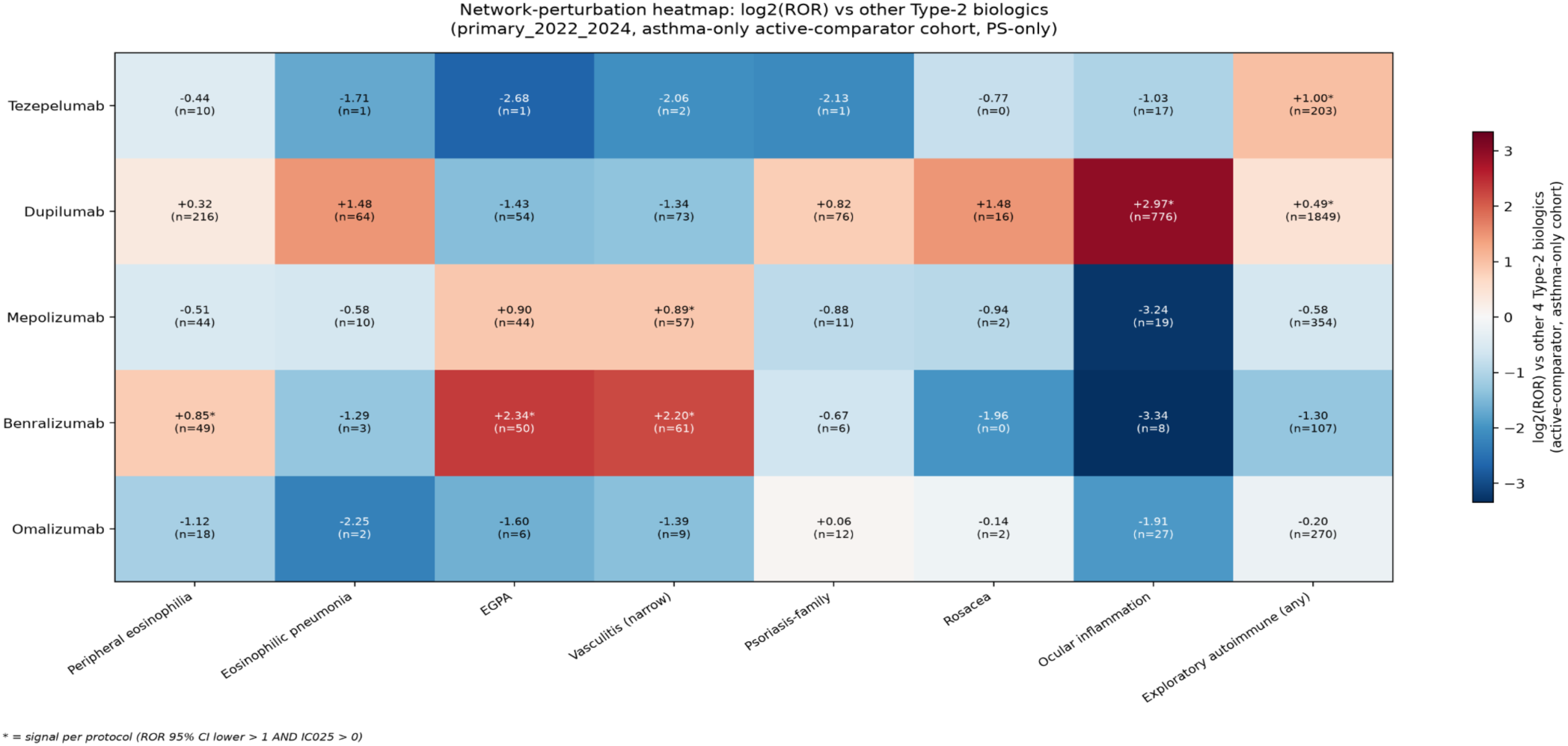
Network-perturbation heatmap of active-comparator disproportionality. log^2^(reporting odds ratio [ROR]) for each of the five biologics against the pooled other four (active-comparator design, 2022–2024 asthma-only cohort), across seven pre-specified event clusters plus one exploratory autoimmune composite (not part of the primary hypothesis set; see Sections 2.3 and 4.4). Warm colors indicate disproportionately increased reporting; cool colors indicate disproportionately decreased reporting relative to the pooled comparator. Asterisks denote a signal that is significant under Benjamini-Hochberg false discovery rate correction (q<0.05, applied jointly across all 40 cells of this matrix) and has ROR>1; this is a stricter, matrix-wide-corrected complement to the unadjusted, per-domain robust-signal criterion used to classify signals in Table 2 (ROR 95% CI lower bound >1 and IC^025^>0 or EBGM^05^>2). The two criteria are concordant for 32 of 35 primary-domain cells; the three exceptions (mepolizumab-EGPA, dupilumab-eosinophilic pneumonia, dupilumab-psoriasis-family) are noted in the Table 2 footnote. n = case count for that cell (the “a” cell of the underlying 2×2 table).

Time-to-onset (event date minus therapy start date, restricted to cases with day-level resolution on both dates) and seriousness (any of FDA’s seven standard seriousness criteria) were also extracted for all qualifying cases.

### 2.4 Statistical analysis

For each drug-event pair, a 2×2 contingency table was constructed comparing the target biologic with the pooled other four (Analysis A) or with the entire FAERS database (Analysis B). We calculated the reporting odds ratio (ROR) with 95% confidence interval (Wald method, Haldane-Anscombe continuity correction when needed), the information component (IC, Bayesian Confidence Propagation Neural Network formula) and its lower 95% credible limit (IC025), and the empirical Bayes geometric mean (EBGM) and its 5th-percentile lower bound (EBGM05), estimated via a Multi-item Gamma Poisson Shrinker fitted across the drug×event matrix of the analysis population.

A drug-event pair was classified as a pre-specified robust signal if it met both the ROR criterion (95% CI lower bound >1) and at least one Bayesian corroboration criterion (IC025 >0 or EBGM05 >2), with a minimum of three reports, consistent with recommended practice that a disproportionality signal should not rest on a frequentist point estimate alone. This criterion was applied, unadjusted for multiplicity, to the seven pre-specified outcomes. As a complementary, more conservative check, Benjamini-Hochberg false discovery rate (FDR) correction (q<0.05, applied jointly across all 40 cells of the primary-domain-by-biologic matrix) was used to construct the network-perturbation heatmap (Figure 2) and to flag exploratory Preferred-Term-level findings. Where the FDR-corrected and pre-specified criteria diverged, both results are reported and the divergence is explicitly flagged (Table 3).

**Table 3.** Analysis A: active-comparator disproportionality results, five biologics × seven pre-specified outcome measures, 2022–2024.

| Domain | Biologic | n cases / N drug | ROR (95% CI) | IC (IC<sub>25</sub>) | EBGM (EBGM<sub>5</sub>) | Robust signal |
| --- | --- | --- | --- | --- | --- | --- |
| **Peripheral eosinophilia** | Tezepelumab | 10 / 2,006 | 0.74 (0.39–1.38) | −0.41 (−1.56) | 0.73 (0.44) | No |
|  | Dupilumab | 216 / 29,652 | 1.25 (1.00–1.56) | 0.12 (−0.16) | 1.08 (0.97) | No |
|  | Mepolizumab | 44 / 8,846 | 0.70 (0.51–0.97) | −0.42 (−0.98) | 0.74 (0.58) | No |
|  | Benralizumab | 49 / 4,372 | 1.80 (1.33–2.44) | 0.73 (0.08) | 1.57 (1.24) | **Yes** |
|  | Omalizumab | 18 / 5,484 | 0.46 (0.29–0.74) | −1.01 (−1.81) | 0.50 (0.34) | No |
| **Eosinophilic pneumonia** | Tezepelumab | 1 / 2,006 | 0.30 (0.04–2.19) | −1.30 (−4.04) | 0.49 (0.18) | No |
|  | Dupilumab | 64 / 29,652 | 2.80 (1.62–4.84) | 0.44 (−0.10) | 1.31 (1.07) | No† |
|  | Mepolizumab | 10 / 8,846 | 0.67 (0.35–1.30) | −0.47 (−1.62) | 0.70 (0.43) | No |
|  | Benralizumab | 3 / 4,372 | 0.41 (0.13–1.30) | −1.09 (−2.92) | 0.50 (0.23) | No |
|  | Omalizumab | 2 / 5,484 | 0.21 (0.05–0.85) | −1.88 (−3.90) | 0.35 (0.15) | No |
| **EGPA** | Tezepelumab | 1 / 2,006 | 0.16 (0.02–1.12) | −2.15 (−4.71) | 0.34 (0.12) | No |
|  | Dupilumab | 54 / 29,652 | 0.37 (0.27–0.52) | −0.75 (−1.24) | 0.59 (0.47) | No |
|  | Mepolizumab | 44 / 8,846 | 1.86 (1.31–2.65) | 0.68 (−0.00) | 1.51 (1.18) | No† |
|  | Benralizumab | 50 / 4,372 | 5.06 (3.60–7.09) | 1.86 (1.00) | 3.54 (3.42) | **Yes** |
|  | Omalizumab | 6 / 5,484 | 0.33 (0.15–0.74) | −1.42 (−2.72) | 0.40 (0.22) | No |
| **Vasculitis (narrow spectrum)** | Tezepelumab | 2 / 2,006 | 0.24 (0.06–0.97) | −1.77 (−3.81) | 0.37 (0.16) | No |
|  | Dupilumab | 73 / 29,652 | 0.39 (0.30–0.52) | −0.70 (−1.12) | 0.61 (0.51) | No |
|  | Mepolizumab | 57 / 8,846 | 1.85 (1.36–2.52) | 0.68 (0.08) | 1.52 (1.23) | **Yes** |
|  | Benralizumab | 61 / 4,372 | 4.60 (3.40–6.22) | 1.77 (1.01) | 3.52 (3.11) | Yes |
|  | Omalizumab | 9 / 5,484 | 0.38 (0.19–0.74) | −1.24 (−2.34) | 0.44 (0.26) | No |
| **Psoriasis-family composite** | Tezepelumab | 1 / 2,006 | 0.23 (0.03–1.64) | −1.65 (−4.30) | 0.42 (0.16) | No |
|  | Dupilumab | 76 / 29,652 | 1.77 (1.16–2.70) | 0.28 (−0.20) | 1.19 (0.98) | No† |
|  | Mepolizumab | 11 / 8,846 | 0.54 (0.29–1.01) | −0.73 (−1.79) | 0.60 (0.37) | No |
|  | Benralizumab | 6 / 4,372 | 0.63 (0.28–1.44) | −0.58 (−2.01) | 0.65 (0.36) | No |
|  | Omalizumab | 12 / 5,484 | 1.04 (0.57–1.91) | 0.05 (−1.09) | 0.95 (0.60) | No |
| **Rosacea** | Tezepelumab | 0 / 2,006 | 0.59 (0.04–9.72) | −1.37 (−6.08) | 0.53 (0.16) | No |
|  | Dupilumab | 16 / 29,652 | 2.79 (0.93–8.36) | 0.43 (−0.64) | 1.20 (0.81) | No |
|  | Mepolizumab | 2 / 8,846 | 0.52 (0.12–2.25) | −0.68 (−2.96) | 0.61 (0.26) | No |
|  | Benralizumab | 0 / 4,372 | 0.26 (0.02–4.24) | −2.16 (−6.58) | 0.43 (0.13) | No |
|  | Omalizumab | 2 / 5,484 | 0.91 (0.21–3.92) | –0.10 (–2.59) | 0.76 (0.32) | No |
| **Ocular inflammation composite** | Tezepelumab | 17 / 2,006 | 0.49 (0.30–0.79) | –0.97 (–1.80) | 0.52 (0.35) | No |
|  | Dupilumab | 776 / 29,652 | 7.81 (6.12–9.97) | 0.64 (0.48) | 1.55 (1.46) | **Yes** |
|  | Mepolizumab | 19 / 8,846 | 0.11 (0.07–0.17) | –2.94 (–3.62) | 0.14 (0.10) | No |
|  | Benralizumab | 8 / 4,372 | 0.10 (0.05–0.20) | –3.12 (–4.15) | 0.13 (0.08) | No |
|  | Omalizumab | 27 / 5,484 | 0.27 (0.18–0.39) | –1.75 (–2.37) | 0.31 (0.22) | No |

ROR = reporting odds ratio; CI = confidence interval; IC = information component (Bayesian Confidence Propagation Neural Network algorithm), shown as point estimate with the lower 95% credible limit (IC<sub>25</sub>) in parentheses; EBGM = empirical Bayes geometric mean, shown as point estimate with its 5th-percentile lower bound (EBGM<sub>5</sub>) in parentheses; EGPA = eosinophilic granulomatosis with polyangiitis. Each biologic is compared against the pooled other four within the asthma-only active-comparator cohort. A signal was classified as robust (“Yes”) if the ROR 95% CI lower bound exceeded 1 and at least one Bayesian criterion (IC<sub>25</sub>>0 or EBGM<sub>5</sub>>2) was met, with a minimum of three reports. †Three cells met the complementary, matrix-wide Benjamini-Hochberg false-discovery-rate-corrected significance threshold ( $q<0.05$ ; Figure 2) despite not meeting the pre-specified, unadjusted robust-signal criterion: mepolizumab-EGPA ( $q=0.0022$ ), dupilumab-eosinophilic pneumonia, and dupilumab-psoriasis-
family. These three associations are reported as exploratory, hypothesis-generating findings (Sections 3.2 and 4.4), not as robust signals.

Time-to-onset distributions were summarized as medians with interquartile ranges and compared between drug groups using the two-sided Mann-Whitney U test (p<0.05). Ten pre-specified sensitivity analyses (Table 2) varied the calendar window, drug-role definition, indication restriction, report seriousness, reporter type, reporting country, and cross-exposure handling. No adjustment for confounding by co-medication, age, sex, or reporting country was performed beyond these design-based restrictions. Analyses were performed in Python 3.12 (pandas, numpy, scipy, statsmodels).

## 3. Results

### 3.1 Study population

Among 4,451,126 deduplicated FAERS reports in the 2022–2024 window across all drugs and indications, 262,191 named one of the five study biologics as Primary Suspect for any indication, and 50,360 met the full asthma-only active-comparator cohort criteria (Table 1; Figure 3): dupilumab (n=29,652, 58.9%), mepolizumab (n=8,846, 17.6%), omalizumab (n=5,484, 10.9%), benralizumab (n=4,372, 8.7%), and tezepelumab (n=2,006, 4.0%). Sex was recorded as female in 29,693 reports (70.3% of the 42,263 reports with known sex), male in 12,570, and unknown, missing, or other in 8,097.

**Figure 3.**
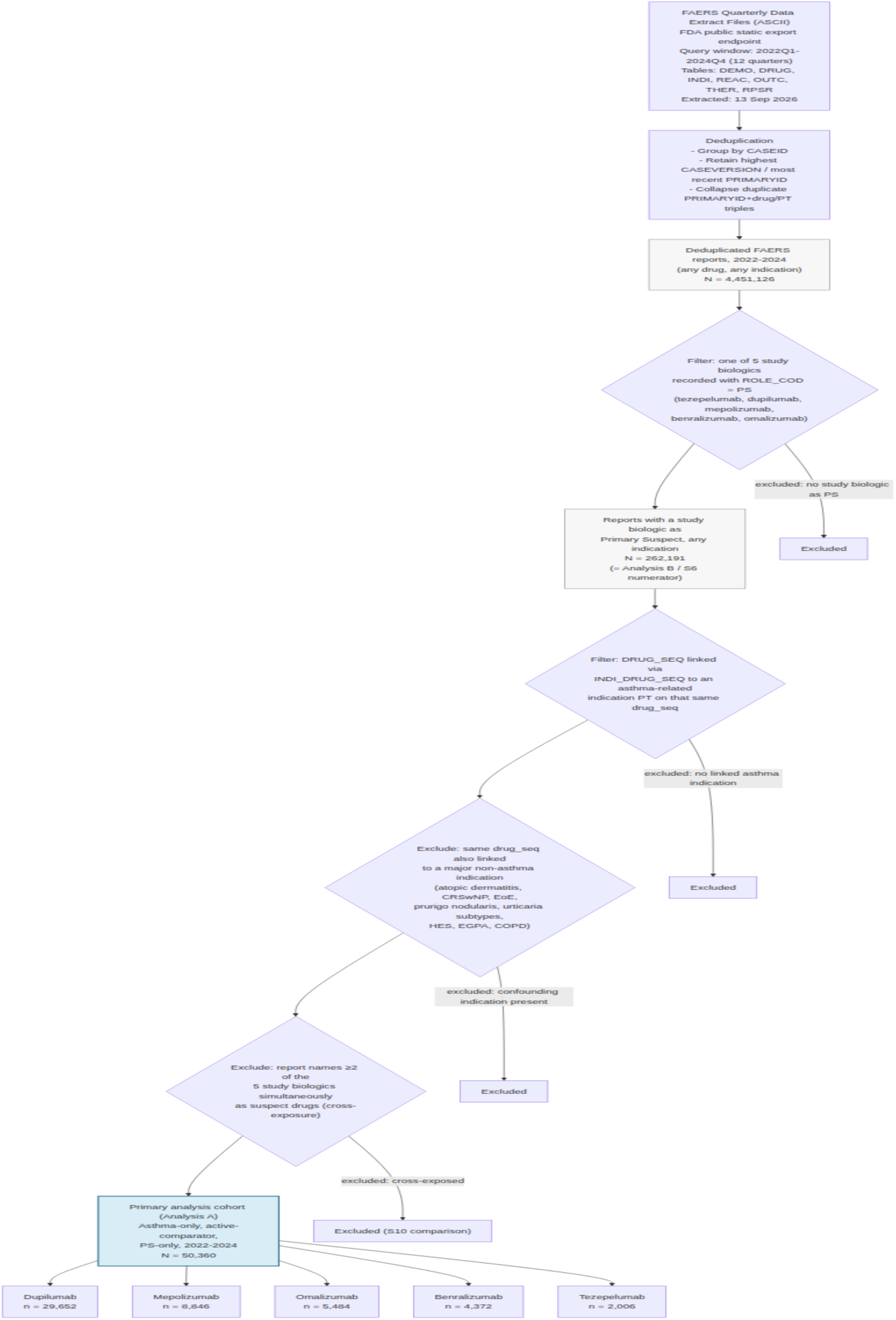
Study design and data extraction flowchart. All 40 quarterly FAERS ASCII extracts (2015Q1–2024Q4) were downloaded directly from FDA’s public static export endpoint on 13 September 2026, with zero data-access failures. The flowchart traces the primary analysis’s 2022–2024 subset from deduplicated reports through role, indication, and cross-exposure filtering to the final 50,360-case active-comparator cohort.

### 3.2 Analysis A: active-comparator disproportionality (primary analysis)

Table 3 and Figure 2 present the full active-comparator disproportionality results for the seven pre-specified outcome measures. Three biologics showed at least one signal meeting the pre-specified robust criterion (ROR 95% CI lower bound >1 and IC025 >0 or EBGM05 >2): benralizumab, mepolizumab, and dupilumab. Omalizumab and tezepelumab met the robust-signal criterion for none of the seven domains.

Benralizumab showed robust signals for all three eosinophilic/vasculitic domains — peripheral eosinophilia (ROR 1.80, 95% CI 1.33–2.44; IC025 +0.08), EGPA (ROR 5.06, 95% CI 3.60–7.09; IC025 +1.00), and narrow-spectrum vasculitis (ROR 4.60, 95% CI 3.40–6.22; IC025 +1.01) — the largest effect sizes observed in the study. Mepolizumab showed a robust vasculitis signal (ROR 1.85, 95% CI 1.36–2.52; IC025 +0.08); its EGPA point estimate was of comparable magnitude (ROR 1.86, 95% CI 1.31–2.65) but the IC025 lower credible limit crossed zero (−0.00), so this specific cell did not meet the pre-specified robust-signal criterion, despite reaching significance under matrix-wide FDR correction (q=0.0022) — a divergence discussed further below.

Dupilumab showed no excess EGPA or vasculitis reporting; both RORs were significantly below 1 (EGPA ROR 0.37, 95% CI 0.27–0.52; vasculitis ROR 0.39, 95% CI 0.30–0.52). Instead, dupilumab showed a large, robust ocular-inflammation signal (ROR 7.81, 95% CI 6.12–9.97; IC025 +0.48; n=776), the single strongest signal in the dataset — dupilumab accounted for 776 of the 847 ocular-inflammation reports in the entire cohort (91.6%). Two further dupilumab associations reached nominal significance on the ROR criterion alone but did not satisfy the pre-specified Bayesian corroboration criterion: eosinophilic pneumonia (ROR 2.80, 95% CI 1.62–4.84; IC025 −0.10, EBGM05 1.07) and the psoriasis-family composite (ROR 1.77, 95% CI 1.16–2.70; IC025 −0.20, EBGM05 0.98). Both were nonetheless significant under matrix-wide FDR correction (q<0.05) and are reported as exploratory, hypothesis-generating associations rather than robust signals; the eosinophilic-pneumonia association additionally attenuated toward the null when Secondary-Suspect reports were included (sensitivity analysis S4, Section 3.7), reinforcing its comparatively fragile evidentiary status relative to the ocular-inflammation signal.

Because dupilumab contributed the large majority of ocular-inflammation reports, the correspondingly low ocular-inflammation RORs computed for the other four biologics against the pooled comparator (omalizumab 0.27, mepolizumab 0.11, benralizumab 0.10, tezepelumab 0.49 [95% CI 0.30–0.79]) primarily reflect this comparator-pool asymmetry rather than an independently protective effect of these agents, since dupilumab itself makes up the great majority of each of their “other four” comparator pools for this specific domain; these values should not be interpreted as evidence of a protective association.

Neither omalizumab nor tezepelumab showed a robust signal in any of the seven domains at the achieved sample size. Tezepelumab’s cohort (n=2,006) yielded wide confidence intervals throughout, including only a single EGPA case (ROR 0.16, 95% CI 0.02–1.12) and a single eosinophilic-pneumonia case (ROR 0.30, 95% CI 0.04–2.19), underscoring the limited precision available for this comparator’s rarer outcomes rather than definitive evidence of safety.

### 3.3 Analysis B: comparison against the whole FAERS background (secondary analysis)

When each biologic’s Primary-Suspect reports (any indication) were instead compared against the entire FAERS database, all five biologics showed an apparent EGPA excess (ROR range 2.8–59.4). A positive signal for every biologic simultaneously is the expected signature of confounding by indication: because all five agents are prescribed predominantly for eosinophilic, T2-high respiratory-allergic disease, each will appear disproportionately associated with a rare eosinophilic diagnosis relative to “all of FAERS,” irrespective of any genuine between-drug difference. This comparison is presented as a methodological contrast rather than a substantive safety finding; the active-comparator design (Analysis A) is treated as primary throughout this manuscript.

### 3.4 Direct pairwise comparisons

Pre-specified direct pairwise comparisons (Supplementary Table S1) corroborated the pattern seen in Analysis A: dupilumab showed significantly higher conjunctivitis reporting than mepolizumab (ROR 8.14) and benralizumab (ROR 9.39), and significantly lower EGPA reporting than both (ROR 0.36 and 0.16, respectively). No pairwise comparison involving tezepelumab reached the robust-signal threshold, reflecting its smaller cohort size rather than a demonstrated absence of effect.

### 3.5 Time-to-onset

Time-to-onset for eosinophilic/vasculitic events differed markedly by biologic (Table 4). Median time-to-onset for EGPA was 20 days after dupilumab (IQR 14–93; 58.3% within 30 days) versus 195 days after benralizumab (IQR 46–243) and 550 days after mepolizumab (IQR 193–895). For narrow-spectrum vasculitis, median time-to-onset was 44 days after dupilumab (IQR 14–118) versus 196 days after benralizumab (IQR 58–222) and 326 days after mepolizumab (IQR 50–841). These differences were statistically significant by two-sided Mann-Whitney U test comparing dupilumab against the pooled mepolizumab-plus-benralizumab group (EGPA: p=0.0016, n=12 vs. 22; vasculitis: p=0.012, n=19 vs. 29). Only a minority of cases in each domain had day-level onset-date resolution sufficient for inclusion in this analysis (for example, 12 of 54 dupilumab EGPA cases, 22%), which limits the precision of these comparisons, even though the direction and significance were consistent across both domains.

**Table 4.** Time-to-onset summary for the primary eosinophilic/vasculitic and ocular domains.

| Biologic | Domain | n | n with TTO data | Median TTO (d) | IQR (d) | %≤30 d | %≤90 d |
| --- | --- | --- | --- | --- | --- | --- | --- |
| Dupilumab | Peripheral eosinophilia | 216 | 44 | 92 | 17–172 | 34.1 | 47.7 |
| Dupilumab | Eosinophilic pneumonia | 64 | 21 | 97 | 46–210 | 9.5 | 42.9 |
| Dupilumab | EGPA | 54 | 12 | **20** | 14–93 | **58.3** | 66.7 |
| Dupilumab | Vasculitis (narrow) | 73 | 19 | **44** | 14–118 | **47.4** | 57.9 |
| Dupilumab | Ocular inflammation | 776 | 95 | 30 | 0–316 | 50.5 | 60.0 |
| Mepolizumab | Peripheral eosinophilia | 44 | 13 | 295 | 57–406 | 15.4 | 30.8 |
| Mepolizumab | EGPA | 44 | 14 | **550** | 193–895 | 7.1 | 21.4 |
| Mepolizumab | Vasculitis (narrow) | 57 | 20 | **326** | 50–841 | 20.0 | 35.0 |
| Benralizumab | Peripheral eosinophilia | 49 | 11 | 222 | 4–285 | 36.4 | 36.4 |
| Benralizumab | EGPA | 50 | 8 | **195** | 46–243 | 25.0 | 37.5 |
| Benralizumab | Vasculitis (narrow) | 61 | 9 | **196** | 58–222 | 22.2 | 33.3 |

*TTO = time-to-onset (event date minus therapy start date), restricted to cases with day-level*
*date resolution on both fields; d = days; IQR = interquartile range; EGPA = eosinophilic*
*granulomatosis with polyangiitis. Two-sided Mann-Whitney U test, dupilumab vs. pooled*
*mepolizumab+benralizumab: EGPA $p=0.0016$ ( $n=12$ vs. $22$ ); vasculitis $p=0.012$ ( $n=19$ vs. $29$ ).*

### 3.6 Seriousness

EGPA and vasculitis reports were classified as serious in 93–100% of cases across all biologics for which sufficient case numbers were available (Table 5), consistent with the expected clinical gravity of these diagnoses irrespective of the implicated drug. Peripheral-eosinophilia reports were also predominantly serious, though less uniformly so (50.9% for dupilumab, 81.6–93.2% for the anti-IL-5-axis agents). By contrast, ocular-inflammation and psoriasis-family reports were considerably less often serious, particularly for dupilumab specifically (ocular inflammation 16.1%; psoriasis-family, approximately 17%), consistent with dupilumab’s ocular/cutaneous signal representing a high-volume but generally non-severe phenomenon, qualitatively distinct from the lower-volume, near-uniformly serious EGPA/vasculitis signal associated with the anti-IL-5-axis agents.

**Table 5.**
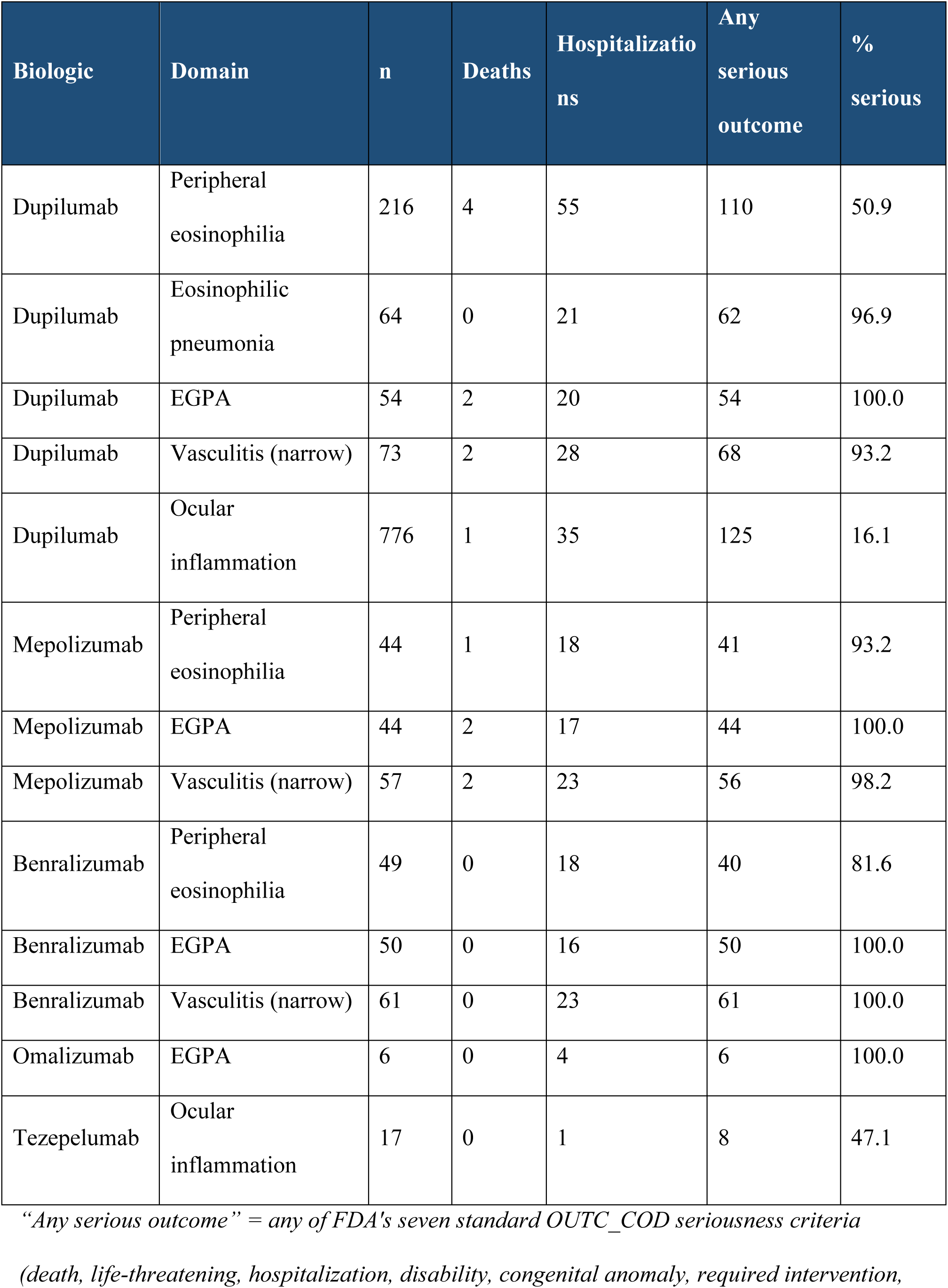
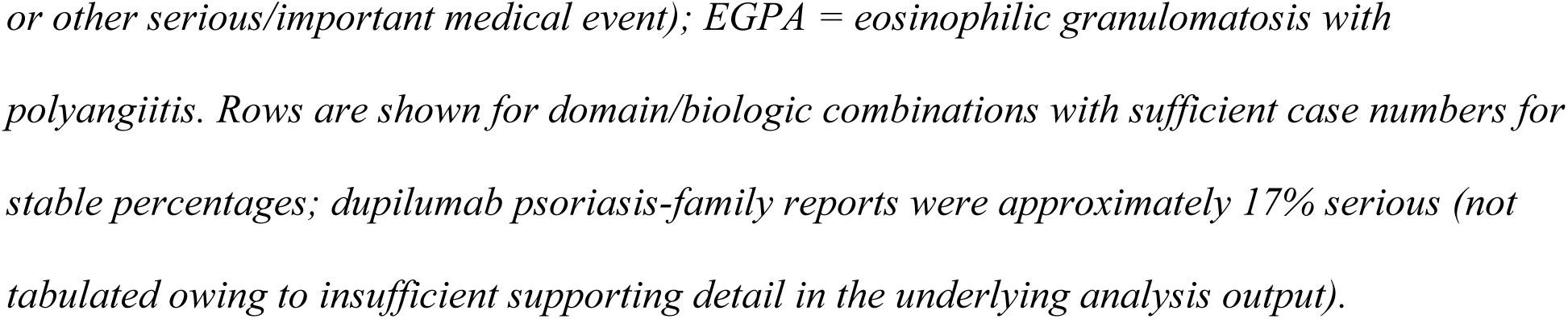
Seriousness summary for the primary domains (representative rows with sufficient case numbers).

### 3.7 Sensitivity analyses

Ten pre-specified sensitivity analyses (Table 2) were performed. The benralizumab EGPA, vasculitis, and peripheral-eosinophilia signals, and the dupilumab ocular-inflammation signal, were robust across role definition, indication restriction, report seriousness, reporter type, and reporting country. No cross-biologic-exposed cases were identified in the primary cohort, so the cross-exposure-exclusion sensitivity analysis (S10) was numerically identical to the primary analysis.

Two findings merit specific emphasis. First, when Secondary-Suspect reports were added to the cohort (S4), dupilumab’s peripheral-eosinophilia and eosinophilic-pneumonia point estimates moved toward or below the null and lost significance, while the benralizumab/mepolizumab EGPA-vasculitis signals and dupilumab’s ocular-inflammation signal persisted — reinforcing that the eosinophilic-pneumonia association is comparatively fragile relative to the robust ocular-inflammation signal. Second, and most importantly, extending the analysis to each drug’s full 2015–2024 post-marketing window (S2) reproduced the benralizumab and dupilumab signals, but mepolizumab’s EGPA and vasculitis signals became null (log₂ ROR +0.38 and +0.28, respectively). Because mepolizumab has been marketed since 2015 — about two years longer than benralizumab and six years longer than tezepelumab — its full post-marketing window spans calendar eras with a systematically different comparator-drug composition than the 2022–2024 primary period. This divergence illustrates how calendar-time confounding can materially alter a disproportionality estimate for an older drug compared against newer comparators, and is the principal reason the common-calendar window, rather than each drug’s full marketing history, was designated primary a priori. A new omalizumab rosacea signal emerged only in this full post-marketing window (ROR 2.52, 95% CI 1.38–4.59; small n) and is noted as hypothesis-generating (Supplementary Figure S2).

## 4. Discussion

### 4.1 Principal findings

In this active-comparator disproportionality analysis of 50,360 asthma-indication FAERS reports, the five T2 asthma biologics did not share a uniform eosinophilic/paradoxical-inflammatory adverse-event signature. Instead, two clinically distinct patterns emerged along mechanistic lines (Figure 1). Benralizumab, and to a lesser and less temporally consistent extent mepolizumab — both targeting the IL-5 axis — were disproportionately associated with EGPA and narrow-spectrum vasculitis, and benralizumab additionally with peripheral eosinophilia. Dupilumab, which blocks the shared IL-4Rα subunit rather than the IL-5 axis, showed no corresponding EGPA/vasculitis excess but instead a large, robust ocular-inflammation signal, accompanied by weaker, less certain associations with eosinophilic pneumonia and paradoxical cutaneous inflammation. Omalizumab and tezepelumab showed no robust composite-domain signal, a genuine negative finding for tezepelumab given its position as the most upstream blocker in the cascade, though one still constrained by its smaller cohort size.

The time-to-onset analysis adds a further, complementary layer to this divergence. EGPA and vasculitis reports clustered much earlier after dupilumab initiation (median 20 and 44 days) than after benralizumab or mepolizumab (median 195–550 days). This pattern is compatible with, though it does not prove, two distinct underlying processes: an early “unmasking” of pre-existing eosinophilic or vasculitic disease during corticosteroid tapering after starting an agent that does not itself deplete eosinophils (dupilumab), versus a later-onset process occurring under sustained, direct IL-5-axis blockade (mepolizumab, benralizumab). This is offered as a plausible hypothesis consistent with the observed temporal pattern, not as a demonstrated mechanism; FAERS records only the reported onset date, not corticosteroid dose, tapering schedule, or pre-treatment eosinophil trajectory.

### 4.2 Biological plausibility

The divergent phenotypes observed here map coherently onto each biologic’s distinct molecular target (Figure 1). Benralizumab depletes eosinophils through both competitive IL-5Rα blockade and antibody-dependent cellular cytotoxicity, producing near-complete peripheral eosinophil depletion; mepolizumab neutralizes circulating IL-5 alone, producing a partial reduction. Insofar as EGPA represents a downstream vasculitic complication of the same eosinophilic-granulomatous process that anti-IL-5-axis agents are designed to treat, a disproportionate reporting association with this end of the eosinophilic spectrum — more pronounced for benralizumab — is biologically plausible, though FAERS data cannot distinguish drug-induced unmasking from detection of pre-existing, subclinical eosinophilic vasculitis in patients whose disease lies on a continuum between severe eosinophilic asthma and EGPA. Dupilumab’s IL-4Rα blockade instead acts on a receptor expressed on conjunctival and periocular epithelium; a disproportionate ocular-surface signal is consistent with this receptor distribution and is, notably, already recognized in dupilumab’s regulatory product labeling (Section 4.3). Omalizumab acts furthest downstream, at the point of IgE-FcεRI engagement, and tezepelumab acts furthest upstream, before the T2 cascade is engaged at all; the absence of a robust signal for either agent is directionally consistent with their positions in the cascade, although absence of signal in spontaneous-report data is not equivalent to demonstrated safety.

### 4.3 Distinguishing expected reactions from emerging pharmacovigilance signals

Ocular surface inflammation is a recognized adverse reaction in dupilumab’s regulatory product labeling; its detection here corroborates an already-known risk, at a magnitude (ROR 7.81) consistent with its prominence in dupilumab’s post-marketing safety profile, rather than representing a novel finding. By contrast, the magnitude of the benralizumab and mepolizumab EGPA/narrow-vasculitis signals under a formal, pre-specified active-comparator design, and dupilumab’s eosinophilic-pneumonia and paradoxical-cutaneous associations, have not, to our knowledge, been systematically quantified using this design in the prior literature. These should be regarded as emerging, hypothesis-generating signals, not confirmed causal associations.

### 4.4 Methodological considerations arising from this analysis

Three methodological points merit explicit discussion. First, the pre-specified unadjusted robust-signal criterion and the matrix-wide FDR-corrected criterion produced discordant classifications for three of 35 primary-domain cells (mepolizumab-EGPA, dupilumab-eosinophilic pneumonia, dupilumab-psoriasis-family): each reached FDR-corrected significance without meeting the stricter, Bayesian-corroborated criterion. We report both classifications and treat all three as exploratory rather than robust; a different pre-specified threshold could reasonably reach a different conclusion for these cells.

Second, active-comparator RORs are only as informative as the comparator pool’s composition: when one biologic dominates a domain’s event counts, as dupilumab does for ocular inflammation (91.6% of cases), the RORs computed for the remaining biologics are mechanically suppressed and should not be read as evidence of a protective effect — a structural property of any active-comparator design, not a flaw specific to this dataset.

Third, EGPA and hypereosinophilic syndrome are both study outcomes and approved indications for anti-IL-5-axis agents, and severe eosinophilic asthma and EGPA lie on a shared biological continuum; although our cohort definition excluded reports with a concurrent EGPA/hypereosinophilic-syndrome indication, FAERS coding cannot fully exclude patients whose asthma was evolving toward, or represented a forme fruste of, EGPA, which should temper causal interpretation of the anti-IL-5-axis EGPA signals specifically. The “exploratory autoimmune (any)” composite in Figure 2 is numerically dominated by non-specific Preferred Terms such as arthralgia and is shown for completeness only, not as evidence of a specific autoimmune signature.

### 4.5 Clinical implications

These findings should be interpreted as hypothesis-generating pharmacovigilance signals relevant to clinicians prescribing and monitoring T2 biologics for severe asthma, not as a basis for changing prescribing practice on their own. The pattern observed supports continued vigilance for EGPA and vasculitic symptoms in patients receiving anti-IL-5-axis therapy, particularly during corticosteroid tapering, and for ocular surface symptoms in patients receiving dupilumab, consistent with existing recommendations for ophthalmologic monitoring during dupilumab therapy. The markedly earlier onset of eosinophilic/vasculitic reports after dupilumab, relative to anti-IL-5-axis agents, may help calibrate monitoring timing, though this derives from a small subset of cases. Because tezepelumab showed no excess signal in any domain, no domain-specific monitoring recommendation is supported for this agent by the present data; this should be read as reassuring but provisional.

### 4.6 Strengths and limitations

This study’s principal strength is its active-comparator design, applied simultaneously across all five currently marketed T2 asthma biologics within a common calendar period, which directly addresses the confounding-by-indication problem that limits conventional whole-database disproportionality analyses (Section 3.3). The pre-specified, dual-criterion robust-signal definition, the ten pre-specified sensitivity analyses, and the explicit comparison between the common-calendar and full post-marketing windows further strengthen confidence in the signals reported as robust.

This study also has several limitations inherent to spontaneous-report disproportionality analysis. First, and most fundamentally, disproportionality metrics quantify relative reporting frequency and cannot, by themselves, establish causality or estimate incidence, because FAERS lacks a denominator of total drug exposure and is subject to substantial, unquantifiable under-reporting. Second, reporting is voluntary and subject to stimulated-reporting and notoriety bias, whereby a drug’s novelty or known safety concerns can disproportionately increase reporting independent of any true change in risk; conversely, long-established drugs such as omalizumab (marketed since 2003) may be under-represented for reasons unrelated to true safety, which may partly explain its muted signal profile here. Third, confounding by indication was addressed by design but cannot be fully excluded: disease severity and treatment history differ systematically across biologics, and the EGPA/eosinophilic-asthma continuum represents a specific residual form of this confounding. Fourth, no case-by-case causality-assessment review (e.g., WHO-UMC, Naranjo) was performed; the signals reported here are hypothesis-generating and require validation through case-level review or comparative pharmacoepidemiological designs before informing a regulatory conclusion. Additional limitations include the EBGM matrix being scoped to the five study biologics rather than the full FAERS database, residual duplicate reporting under distinct case identifiers, and approximated (rather than licensed) MedDRA query definitions for the eosinophilic-pneumonia and vasculitis domains.

## 5. Conclusion

In this FAERS active-comparator disproportionality analysis of 50,360 asthma-indication reports, the five Type 2 asthma biologics showed mechanistically coherent, drug-specific — not class-uniform — patterns of eosinophilic and paradoxical inflammatory adverse-event reporting. Benralizumab showed the most consistent and largest-magnitude signals, spanning peripheral eosinophilia, EGPA, and vasculitis; mepolizumab showed a robust vasculitis signal but an EGPA association that did not meet the full robustness threshold and did not replicate over the full post-marketing window, a discordance attributable to calendar-time confounding rather than a genuine absence of association. Dupilumab showed a robust, large-magnitude ocular-inflammation signal with markedly earlier onset than the anti-IL-5-axis agents’ eosinophilic/vasculitic events. Tezepelumab and omalizumab showed no robust signal in any domain. These findings support targeted, mechanism-informed adverse-event monitoring rather than a uniform, class-wide vigilance strategy, while underscoring that disproportionality signals from spontaneous reports are hypothesis-generating and require independent confirmation before informing causal or regulatory conclusions.

## Data Availability

The FAERS quarterly data extract files analyzed are publicly available at no cost from the FDA (https://fis.fda.gov/extensions/FPD-QDE-FAERS/FPD-QDE-FAERS.html). The analysis pipeline (Python 3.12; pandas, numpy, scipy, statsmodels, matplotlib, seaborn) is available from the corresponding author upon reasonable request.

## Declarations

## Acknowledgments

None.

## Ethics approval

This study used publicly available, de-identified, aggregate spontaneous adverse-event report data (FAERS) and did not involve identifiable patient information or direct patient contact; such analyses are generally exempt from formal institutional ethics committee review, consistent with standard practice for aggregate pharmacovigilance database research.

## Declaration of Generative AI Use

During the preparation of this manuscript, the authors utilized Claude Sonnet 5 (Anthropic) solely for grammatical editing, syntax verification, and structural formatting. The authors performed extensive line-by-line manual verification, retain full responsibility for the data interpretation and intellectual content, and affirm that no artificial intelligence software was utilized for raw data processing or statistical computation.

