## Supplementary Tables and Figures for "Divergent Eosinophilic and Paradoxical Inflammatory Adverse-Event Signatures Across Five Type 2 Asthma Biologics: A FAERS Active-Comparator Disproportionality Analysis"

### Supplementary Material

To conform to this journal's 8-display-item limit for the main text, two supportive display items are provided here rather than in the main manuscript:
Supplementary Table S1 (pre-specified direct pairwise comparisons among biologics, referenced in Sections 2.1 and 3.4) and Supplementary Figure S1 (within-drug distribution of the seven pre-specified adverse-event domains).

Supplementary Figure S2 presents the network-perturbation heatmap for the secondary, full 2015–2024 post-marketing window (sensitivity analysis S2), constructed identically to Figure 2 but using each drug's complete marketing history through December 2024 rather than the common-calendar 2022–2024 primary period, and is provided to support the primary-versus-full-window comparison discussed in Sections 3.7 and 4.4 (in particular, the loss of mepolizumab's EGPA/vasculitis signal over the longer window); the 2022–2024 common-calendar analysis (Figure 2) remains the pre-specified primary comparison. Individual Preferred-Term-level results underlying the exploratory autoimmune composite, and the full per-cell case counts and Benjamini-Hochberg q-values underlying Figure 2 and Supplementary Figure S2, are available from the corresponding author upon reasonable request.

**Supplementary Table S1. Pre-specified direct pairwise comparisons among biologics, 2022–2024.**

| Comparison | Endpoint | n (target) | n (comp.) | ROR | 95% CI | Robust signal |
|---|---|---|---|---|---|---|
| Dupilumab vs. tezepelumab | Eosinophilia | 78 | 2 | 2.64 | 0.65–10.76 | No |
| Dupilumab vs. tezepelumab | EGPA | 54 | 1 | 3.66 | 0.51–26.46 | No |
| Dupilumab vs. tezepelumab | Eosinophilic pneumonia | 60 | 1 | 4.07 | 0.56–29.35 | No |
| Dupilumab vs. tezepelumab | Psoriasis | 76 | 1 | 5.15 | 0.72–37.07 | No |
| Dupilumab vs. tezepelumab | Rosacea | 16 | 0 | 2.23 | 0.13–37.25 | No |
| Dupilumab vs. tezepelumab | Conjunctivitis | 190 | 12 | 1.07 | 0.60–1.92 | No |
| Dupilumab vs. mepolizumab | Eosinophilia | 78 | 6 | **3.89** | **1.69–8.92** | **Yes** |
| Dupilumab vs. mepolizumab | EGPA | 54 | 44 | 0.36 | 0.24–0.54 | No |
| Dupilumab vs. mepolizumab | Eosinophilic pneumonia | 60 | 8 | **2.24** | **1.07–4.69** | **Yes** |
| Dupilumab vs. mepolizumab | Psoriasis | 76 | 11 | **2.06** | **1.10–3.89** | **Yes** |

| Comparison | Endpoint | n (target) | n (comp.) | ROR | 95% CI | Robust signal |
|---|---|---|---|---|---|---|
| Dupilumab vs. mepolizumab | Rosacea | 16 | 2 | 2.39 | 0.55–10.38 | No |
| Dupilumab vs. mepolizumab | Conjunctivitis | 190 | 7 | **8.14** | **3.83–17.32** | **Yes** |
| Dupilumab vs. benralizumab | Eosinophilia | 78 | 8 | 1.44 | 0.69–2.98 | No |
| Dupilumab vs. benralizumab | EGPA | 54 | 50 | 0.16 | 0.11–0.23 | No |
| Dupilumab vs. benralizumab | Eosinophilic pneumonia | 60 | 3 | 2.95 | 0.93–9.42 | No |
| Dupilumab vs. benralizumab | Psoriasis | 76 | 6 | 1.87 | 0.81–4.30 | No |
| Dupilumab vs. benralizumab | Rosacea | 16 | 0 | 4.87 | 0.29–81.17 | No |
| Dupilumab vs. benralizumab | Conjunctivitis | 190 | 3 | **9.39** | **3.00–29.39** | **Yes** |
| Tezepelumab vs. pooled T2-axis* | Eosinophilia | 2 | 92 | 0.46 | 0.11–1.88 | No |
| Tezepelumab vs. pooled T2-axis* | EGPA | 1 | 148 | 0.14 | 0.02–1.03 | No |
| Tezepelumab vs. pooled T2-axis* | Eosinophilic pneumonia | 1 | 71 | 0.30 | 0.04–2.16 | No |

| Comparison | Endpoint | n (target) | n (comp.) | ROR | 95% CI | Robust signal |
|---|---|---|---|---|---|---|
| Tezepelumab vs. pooled T2-axis* | Psoriasis | 1 | 93 | 0.23 | 0.03–1.65 | No |
| Tezepelumab vs. pooled T2-axis* | Rosacea | 0 | 18 | 0.58 | 0.03–9.58 | No |
| Tezepelumab vs. pooled T2-axis* | Conjunctivitis | 12 | 200 | 1.28 | 0.72–2.30 | No |

*ROR = reporting odds ratio; CI = confidence interval; EGPA = eosinophilic granulomatosis with polyangiitis; T2 = type 2. Endpoints in this table use single, specific MedDRA Preferred Terms (eosinophilia, EGPA, eosinophilic pneumonia, psoriasis, rosacea, conjunctivitis), per the pre-specified pairwise-comparison protocol, rather than the composite domains used in Table 2; case counts therefore differ slightly from the composite totals reported there.*

**Pooled T2-axis comparator = dupilumab + mepolizumab + benralizumab (omalizumab excluded, as pre-specified, because anti-IgE blockade is not part of the upstream/downstream cytokine-receptor cascade shared by the other three agents).*

**Supplementary Figure S1. Within-drug distribution of the seven pre-specified adverse-event domains.**

Percentage of each biologic's own asthma-cohort reports falling within each pre-specified adverse-event domain, shown as a clustered bar chart with case counts (n) annotated above each bar. This figure is complementary to Figure 2: it displays absolute within-drug reporting prevalence rather than between-drug disproportionality, and should not be used to infer relative risk between biologics.

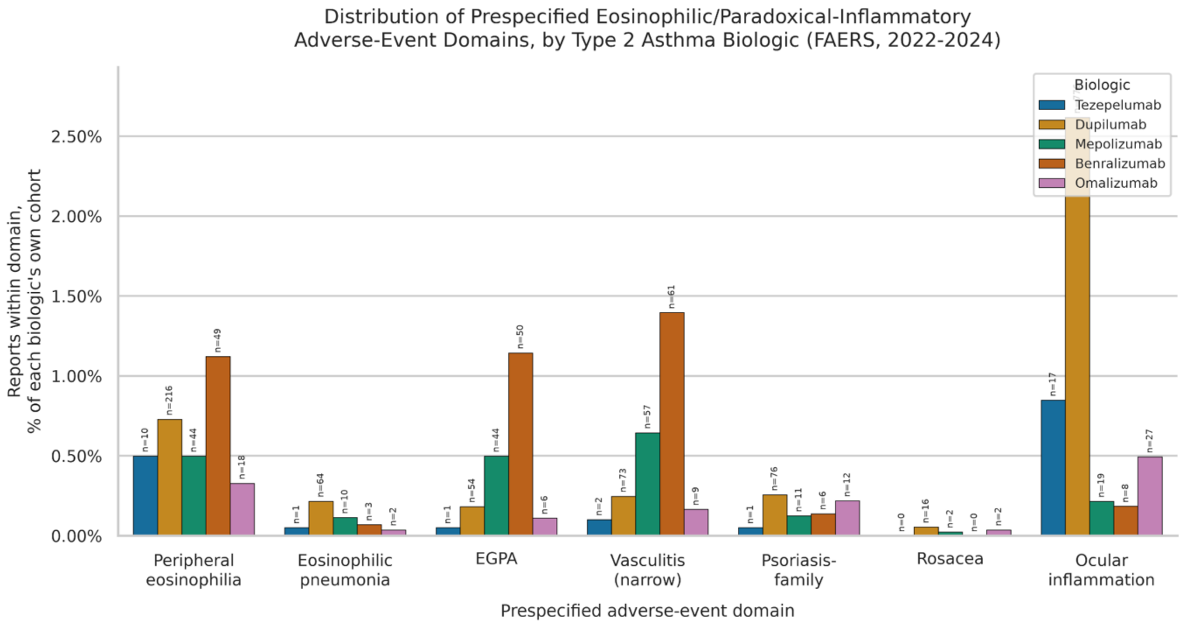

**Supplementary Figure S2. Network-perturbation heatmap of active-comparator disproportionality over the full 2015–2024 post-marketing window (sensitivity analysis S2).**

log$^2$(ROR) for each of the five biologics against the pooled other four, using each drug's complete post-marketing exposure through December 2024 rather than the 2022–2024 common-calendar primary period. Asterisks denote a signal significant under Benjamini-Hochberg false discovery rate correction ($q<0.05$, applied across all 40 cells of this matrix) with ROR>1, as in Figure 2. Note the loss of mepolizumab's EGPA and vasculitis signals over this longer window relative to the primary analysis (Figure 2), attributable to calendar-time confounding (Section 4.4), and the persistence of the benralizumab EGPA/vasculitis/peripheral-eosinophilia and dupilumab ocular-inflammation signals across both windows.

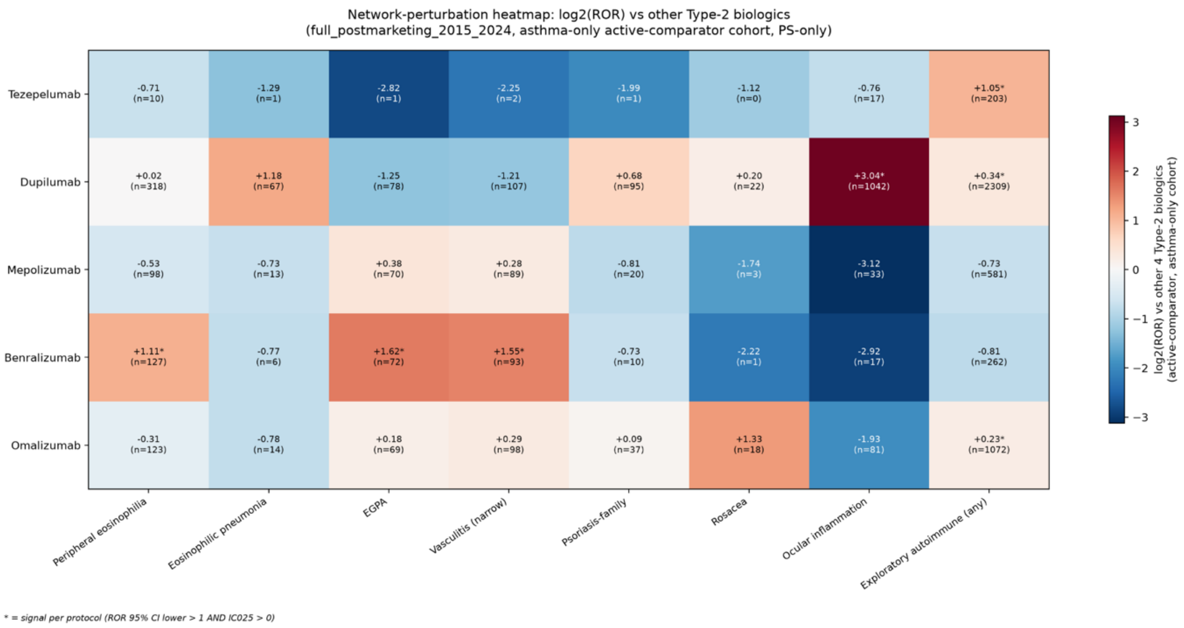
